# Enhanced Recovery After Surgery (ERAS) Increases Long-Term Survival Rate after Surgery in Colorectal Cancer Patients: A Systematic Review and Meta-Analysis

**DOI:** 10.64898/2026.03.05.26347672

**Authors:** Kanglu Yang, Xiaorong Liu, Jianyong Cui, Zhiwei Liu, Jin Liu, Jiyue Zhang, Yonge Wu, Haibin Ji, Qiangpu Chen

## Abstract

**Background:** Enhanced Recovery After Surgery (ERAS) optimizes perioperative management for colorectal cancer (CRC), improving short-term outcomes, but its impact on long-term outcomes remains inconclusive, supporting the need for this meta-analysis. This study evaluates the effect of perioperative ERAS (therapy-focused) on 1-, 2-, 3-, and 5-year postoperative survival in patients with CRC.

**Methods:** We conducted a systematic review and meta-analysis following a pre-registered protocol in accordance with Preferred Reporting Items for Systematic Reviews and Meta-Analyses guidelines. PubMed, Web of Science, Embase, Medline Ovid, and Cochrane Library Wiley were searched up to December 31, 2025, for clinical studies reporting long-term postoperative survival outcomes of patients with CRC undergoing ERAS implementation. Of 1,063 retrieved reports, 10 studies (5,876 patients) were included in Kaplan-Meier-based meta-analyses and eight studies (5,556 patients) in aggregated data meta-analyses. Data extraction was performed independently by two reviewers, with study quality and risk of bias assessed using the Newcastle-Ottawa Scale (NOS) and RevMan software. Effect sizes were pooled using fixed- or random-effects models according to heterogeneity, with cross-validation and subgroup analyses examining the influence of tumor stage and ERAS adherence. The pre-specified primary outcome was postoperative overall survival (OS) ≥12 months, and the secondary outcome was disease-free survival (DFS).

**Results:** ERAS significantly improved OS at 1 year (93.2%, 95% CI: 92.3-94.2 vs. 90.2%, 95% CI: 89.1-91.2), 2 years (86.7% vs. 81.3%), 3 years (81.1% vs. 72.4%), 5 years (70.9% vs. 60.6%) (all P<0.01). The pooled HR for mortality was 0.72 (95% CI: 0.63-0.83, P<0.01), indicating a 28% reduction in long-term mortality. Stage I-II tumors and ERAS adherence ≥70% conferred the greatest benefits. DFS did not show a statistically significant improvement (HR=0.90, 95% CI: 0.68-1.19, P=0.45). Included studies were of moderate to high quality (NOS score 6-9).

**Conclusions:** Perioperative ERAS significantly improves 1- to 5-year OS and reduces long-term mortality in patients with CRC, with the greatest benefits in early-stage disease and high adherence. These findings support ERAS as a critical component of comprehensive CRC care.

## Introduction

Colorectal cancer (CRC) is the third most common malignancy worldwide and a leading cause of cancer death, with surgery as the primary treatment.^1, 2^ Enhanced Recovery After Surgery (ERAS) is a multidisciplinary perioperative care approach that minimizes pathophysiological stress through preoperative prehabilitation, precise intraoperative management, and early postoperative rehabilitation, promoting faster organ recovery.^3^

Substantial evidence demonstrates ERAS in CRC surgery improves outcomes, reducing postoperative complications, hospital stays, and costs.^4,5^ The European Society for Clinical Nutrition and Metabolism strongly recommends ERAS for curative or palliative CRC surgery.^6–8^ Although ERAS has clear short-term benefits, its impact on long-term survival remains controversial: some studies report improved nutritional status and reduced inflammation that may support survival, while others show no prognostic benefit. ^9–11^ This study systematically reviews literature since ERAS inception. Using Kaplan-Meier (KM) reconstruction and pooled analysis, we assess subgroup effects of tumor stage and ERAS adherence to clarify ERAS’s role in long-term CRC management and guide individualized implementation.

## Materials and Methods

This systematic review and meta-analysis followed the Preferred Reporting Items for Systematic Reviews and Meta-Analyses (PRISMA) 2020 guidelines for data collection and reporting.^12^ The protocol was pre-registered on the international systematic review prospective registration platform PROSPERO (CRD42024618247).

### Search Strategy

We searched PubMed, Embase, Cochrane Library, and Web of Science databases (January 1, 1995, to December 31, 2025), covering key stages of ERAS in colorectal surgery. Medical Subject Headings (MeSH) terms included “Enhanced Recovery After Surgery,” “Colorectal Neoplasm,” “Colorectal Cancer Surgery,” along with “Survival,” “Mortality,” and “Survival Analysis.” Keywords encompassed ERAS-related terms: Enhanced Recovery After Surgery, Fast-track surgery, ERAS protocol; disease-related: colorectal cancer, colorectal neoplasm; outcome-related: survival rate, overall survival (OS), disease-free survival (DFS), long-term outcome. MeSH terms and keywords were combined using Boolean operators “AND”/“OR,” with no language restrictions. Two researchers (Zhang, Liu) independently conducted reverse citation tracking and used “similar articles” searches to ensure comprehensive coverage.^13^

### Inclusion and Exclusion Criteria

We included randomized controlled trials (RCTs) and prospective/retrospective cohort studies in English with a defined control group receiving conventional perioperative care. Eligible studies enrolled adults (≥18 years) undergoing CRC surgery, regardless of TNM stage, without severe comorbidities (e.g., severe cardiac, hepatic, or renal failure). The experimental group implemented an ERAS protocol with ≥5 core components (e.g., optimized preoperative fasting, multimodal analgesia, early oral intake/mobility) and the control group received conventional perioperative care (e.g., standard fasting, single-modality analgesia, delayed mobility). Primary outcomes were OS ≥12 months post-surgery; secondary outcomes included DFS, tumor-stage effects, and the impact of ERAS adherence (≥70% as high) on survival. Studies required complete data for meta-analysis (KM curves with person-years or hazard ratios [HR] with 95% confidence intervals [CI]).

We excluded non-controlled studies (e.g., commentaries, reviews, case reports, study protocols, single-arm cohort studies); studies with a single-group sample size <20 cases, due to increased risk of bias in small-sample studies; research involving patients undergoing surgery for non-CRC (e.g., gastric or pancreatic cancer) or with concomitant malignancies, studies lacking postoperative follow-up survival data (e.g., reporting only perioperative complications) or with survival data that could not be extracted from KM curves/text (e.g., missing person-years at risk or HRs); and non-English literature without official/published English translations, as data extraction from such sources is prone to errors).

### Study Selection and Data Extraction

A double-blind independent screening process was used. Two researchers (Yang and Wu) first screened titles and abstracts to exclude ineligible studies, followed by a full-text review for final inclusion. Disagreements were resolved by discussion, with third-party adjudication (Cui) when necessary. Pre-screening calibration standards were applied, and screening consistency was assessed using Cohen’s kappa coefficient (κ > 0.75 indicating good agreement).^14^ Two researchers (Cui and Liu) independently extracted data using a pre-designed form, including study details (first author, publication year, country, study type), participant characteristics (sample size, age, tumor stage, surgical approach), intervention specifics (ERAS modules, compliance criteria), outcomes (1/2/3/5-year OS/DFS, HR with 95% CI, KM data), and quality assessment-related information. Extracted data was cross-checked, maintaining an error rate ≤5%.

### Quality Appraisal of Included Studies

Sleep quality was assessed using differentiated tools. RCTs were evaluated with the Cochrane Risk of Bias tool (RoB 2.0) across six domains: random sequence generation, allocation concealment, blinding, completeness of outcome data, selective reporting, and other biases. Cohort studies were assessed using the Newcastle-Ottawa Scale (NOS) across three dimensions: participant selection (4 points), group comparability (2 points), and outcome measurement (3 points), with scores ≥6 points indicating moderate-to-high quality.^15^ Two researchers (Cui and Liu) independently conducted assessments, resolving disagreements by discussion. Risk-of-bias plots were generated using RevMan 5.4 (Cochrane Collaboration, Copenhagen, Denmark).

### Statistical Analysis

Individual Patient Data are ideal for unbiased effect-size pooling and subgroup analysis, but are difficult to obtain due to privacy and data-sharing restrictions. Aggregated Data often lack critical survival information, creating a methodological bottleneck for survival meta-analyses. To address this, we reconstructed survival data from published KM curves, a method validated in prior analyses.^16,17^ Extracted data included study details, baseline characteristics, and survival outcome. KM curve extraction followed a two-step process: first, Origin Pro 2024 software (OriginLab, Northampton, USA) digitized KM curves using the “Curve Point Extraction Tool,” and at least 10 key nodes (start, inflection, and end) were recorded per curve to ensure accuracy.^18^

Subsequently, the digitized KM coordinates were imported into the Shiny 1.2.2.0.35 web application (R-based) developed by Liu et al., which integrates Guyot et al.’s iterative algorithm to reverse-engineer individual survival data (Reconstructed Patient Data) from three-dimensional time-survival-at-risk data.^19,20^ Data accuracy was validated using a triple-check approach: (1) reconstructed survival probabilities were compared with reported rates (e.g., 5-year survival), allowing error ≤3%; (2) two researchers (Cui and Liu) independently plotted reconstructed KM curves, with overlap assessed visually and quantified via Intraclass Correlation Coefficient (ICC) (ICC ≥ 0.9 considered valid);^21,22^ (3) ambiguous survival data prompted requests to corresponding authors, and studies without responses were excluded.

KM-based meta-analysis and pooled data meta-analysis were conducted concurrently to cross-validate results. Pooled analysis HRs with 95% CI, with HR<1 indicating lower long-term mortality in the ERAS group. All analyses were performed in RevMan 5.4 software, with P<0.05 considered statistically significant for two-tailed tests.^23^

Heterogeneity was assessed using Cochran’s Q test (α=0.10) and the I² index: I²<25% indicated low, 25%≤I²≤50% moderate, and I²>50% high heterogeneity. Random-effects models were applied if I² > 50%; otherwise, fixed-effects models were used. Sources of heterogeneity were explored via pre-specified subgroup and sensitivity analyses: (1) pooling studies reporting DFS; (2) stratifying by tumor stage (Stage I-II vs. Stage III-IV) per AJCC 8th edition criteria;^24^ (3) stratifying by ERAS compliance (high ≥70% vs low <70%) per ERAS Society 2024 Guidelines.^25^ Sensitivity analysis employed a leave-one-out approach; pooled HRs with 95% CI unchanged after exclusions were considered robust.

## Results

The search and screening process is illustrated in Figure 1 (PRISMA 2020 flow diagram). Database searches identified 1,063 records. After excluding duplicates and irrelevant entries (n=637), 426 titles and abstracts were screened. Thirteen full-text articles were assessed for eligibility, with three excluded for insufficient data. Ultimately, 10 studies were included in both qualitative and quantitative analyses: all 10 studies (5,867 patients^28–37^) were included in the KM meta-analysis, while eight studies (5,556 patients^28,29,31,32,34–37^) contributed to the pooled-data meta-analysis.

**Figure 1:**
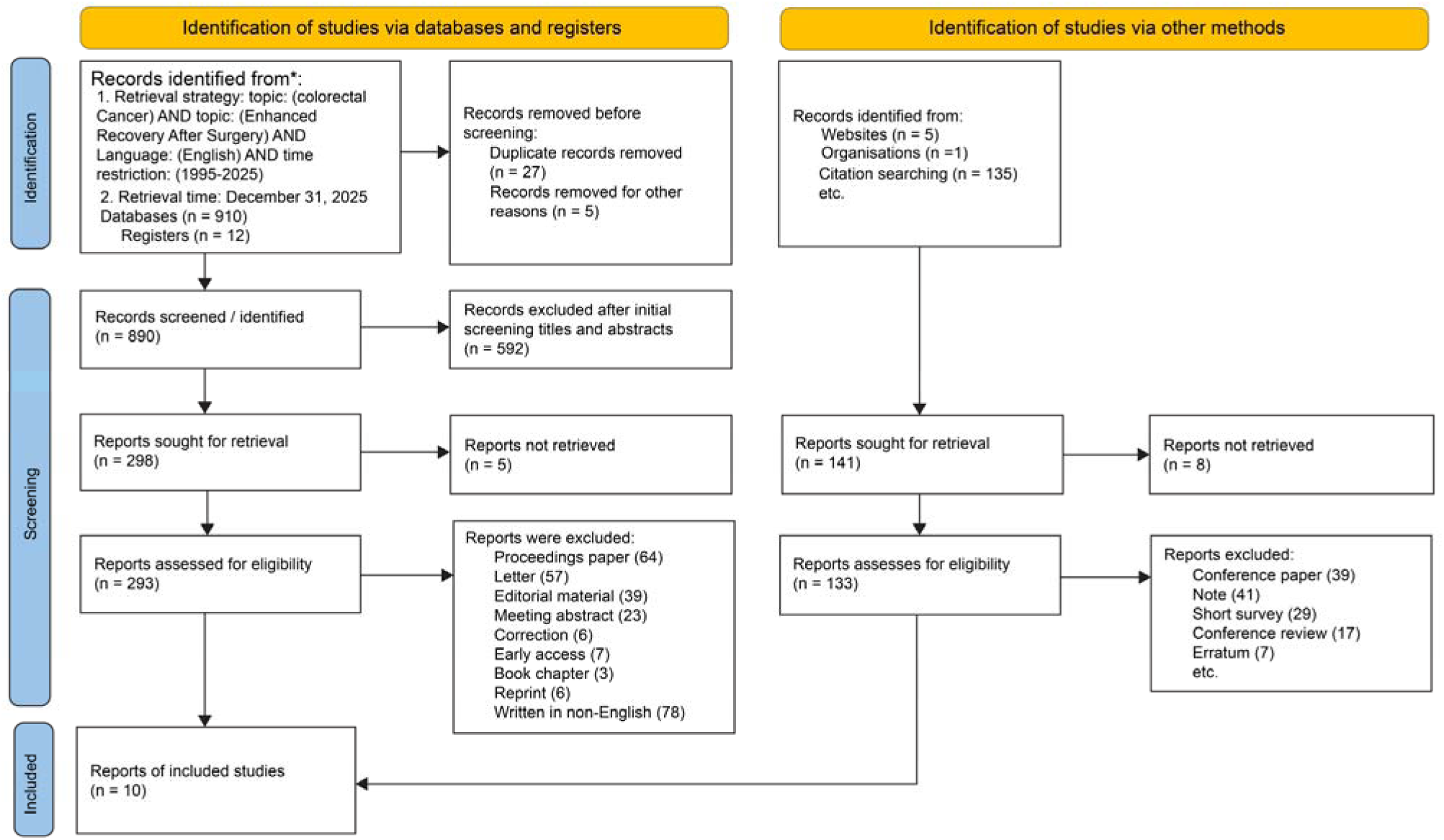
Literature Search and Screening Flowchart

The basic characteristics of included studies are summarized in Tables 1 and 2, detailing location, population, sample size, age, follow-up duration, and meta-analysis type. Ten studies were conducted across three continents: Europe (six studies,^28,30,33,34–36^ 3672 patients), North America (three studies,^29,31,37^ 1,846 patients), and Asia (one study,^32^ 349 patients). All were observational (cohort) studies, mostly conducted after January 1, 2005, and published between 2016 and 2025. Each included an ERAS intervention group and a control group receiving standard perioperative care. Studies were included in either KM-based or pooled data meta-analyses according to data type.

**Table 1:** Basic Characteristics of Included Studies Table 1 presents the sources of included studies, study periods, number of patients in the Enhanced Recovery After Surgery (ERAS) group, age (mean ± standard deviation or median [interquartile range]), median follow-up duration (interquartile range), and whether Kaplan-Meier (KM)-based meta-analysis and pooled data meta-analysis were included. Abbreviations: “KM” denotes Kaplan-Meier; “No” indicates absence of relevant meta-analysis data.

| Source | Study period | No. of patients with ERAS | Age, mean (SD) or median (IQR), y | Follow-up, median(IQR), y | KM-based meta-analysis | Aggregate-data meta-analysis |
| --- | --- | --- | --- | --- | --- | --- |
| F Tidadini et al, 2022, France<sup>36</sup> | 2005-2017 | 497(1001) | 70[61;78] | 3 years | Yes | Yes |
| Andres Zorrilla-Vaca et al, 2022, America<sup>31</sup> | 2010-2016 | 307(646) | 73[63;80] | 5 years | Yes | Yes |
| F Tidadini et al, 2024, France<sup>28</sup> | 2005-2017 | 486(981) | 70[61;78] | 5 years | Yes | Yes |
| Ibrahim Gomaa et al, 2024, America<sup>29</sup> | 2005-2018 | 320(600) | 58.5(13.4) | 10 years | Yes | Yes |
| Jacopo Crippa et al, 2023, Italy<sup>30</sup> | 2014-2015 | 52(105) | 65.48(13) | 5 years | Yes | No |
| Magdalena Pisarska et al, 2025, Poland<sup>35</sup> | 2013-2017 | 340(468) | 65.2(13.5) | 5 years | Yes | Yes |
| P. Viannay et al, 2018, France<sup>33</sup> | 2010-2014 | 52(206) | 70.9(12.1) | 3 years | Yes | No |
| Ulf O. Gustafsson et al, 2016, Sweden<sup>34</sup> | 2002-2007 | 273(911) | 68.7(11.7) | 5 years | Yes | Yes |
| Varut Lohsiriwat et al, 2021, Thailand<sup>32</sup> | 2010-2016 | 70(349) | 62.1(13.8) | 7 years | Yes | Yes |
| Quiram B J et al, 2019, America<sup>37</sup> | 2005-2018 | 320(600) | 58.5(13.4) | 3 years | Yes | Yes |

**Table 2:**
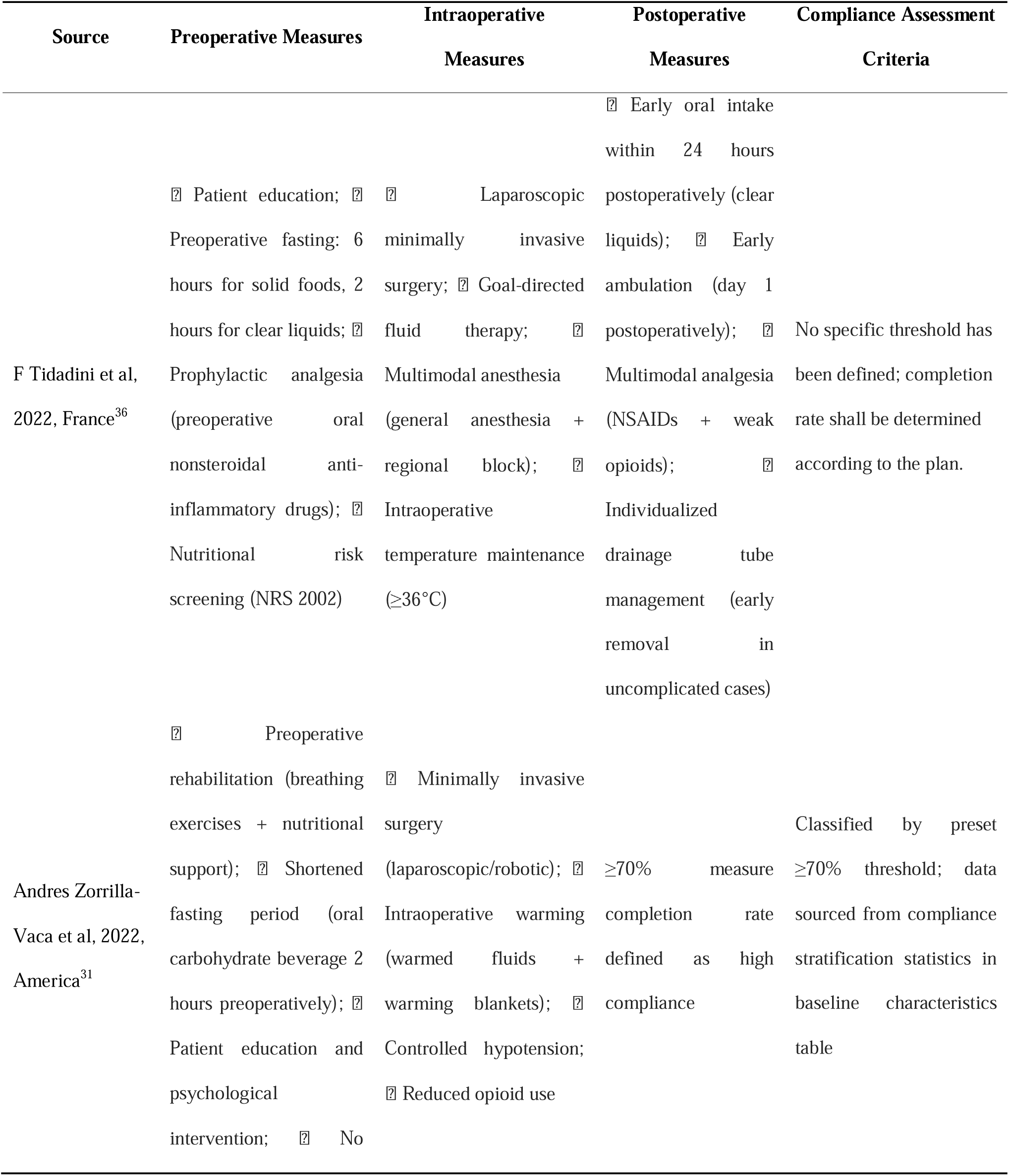
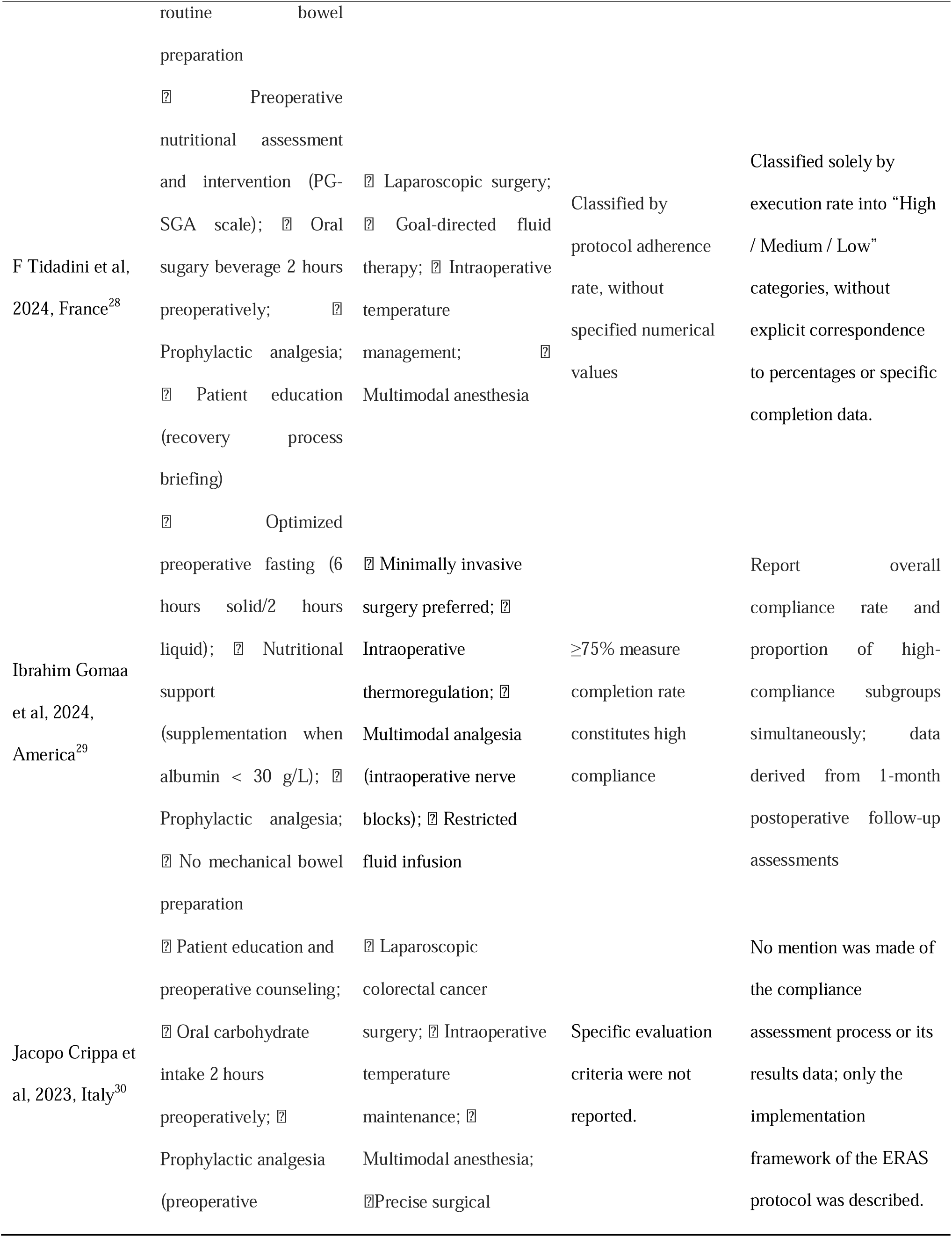
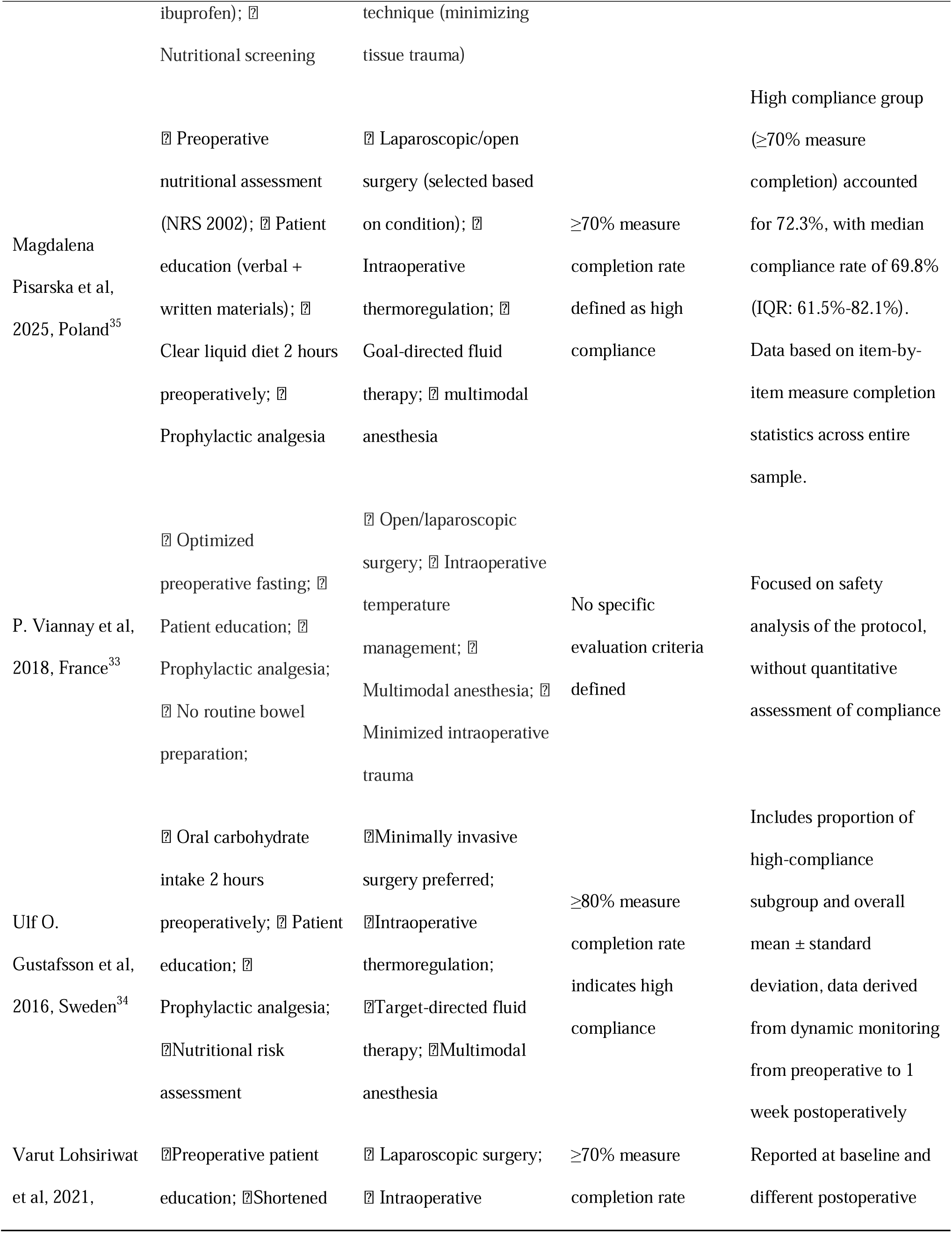
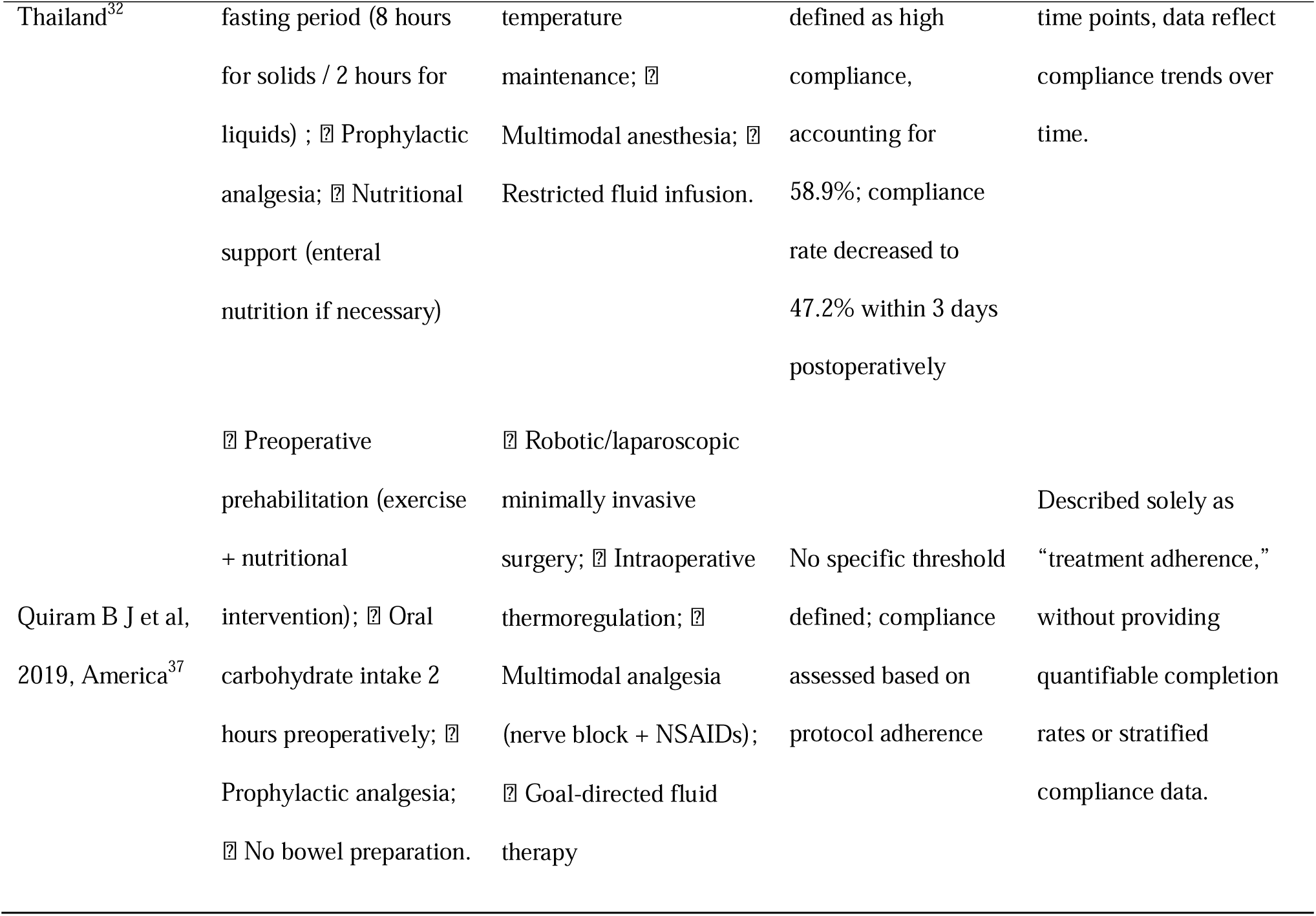
The implementation details and compliance assessment criteria for Enhanced Recovery After Surgery (ERAS) protocols across the 10 included studies. Table 2 details the intervention components (preoperative, intraoperative, postoperative) and compliance assessment criteria included in the studies, categorized according to the ERAS Society’s 2024 Core Module Guidelines for Colorectal Surgery. Compliance data and standards are directly excerpted from the original studies; those without quantified data are labeled as “no specific threshold defined” or “specific assessment criteria not reported.” Terminology (surgical procedures, nutritional assessment tools, etc.) aligns with the original studies, providing a consistent basis for analyzing ERAS protocol heterogeneity and the compliance-outcome relationship.

### Quality of Studies

All included studies were of satisfactory to good quality (Table 3). Each achieved ≥1 star for “comparability of groups” (some scored 2), indicating overall high quality and manageable risk of bias.

**Table 3:**
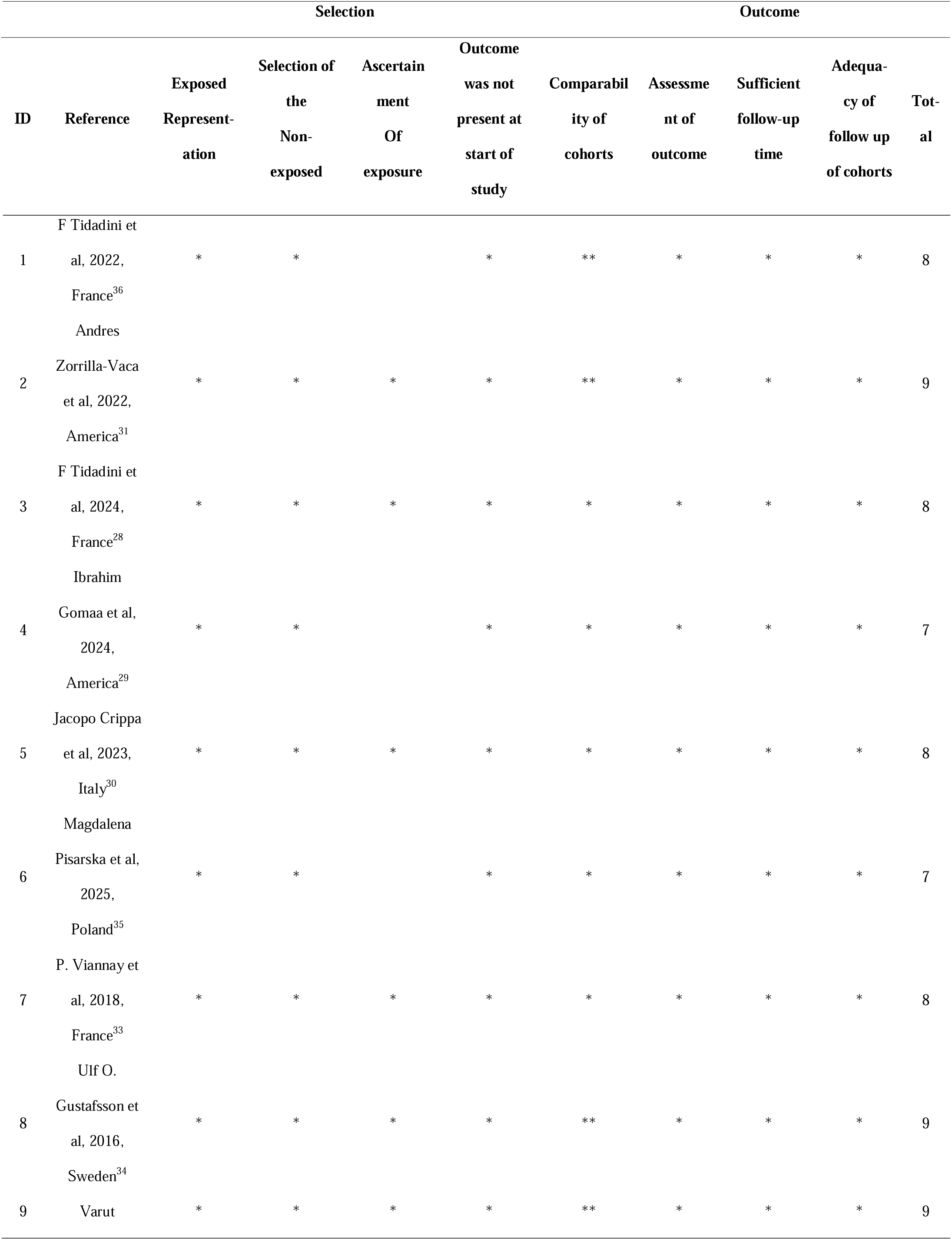
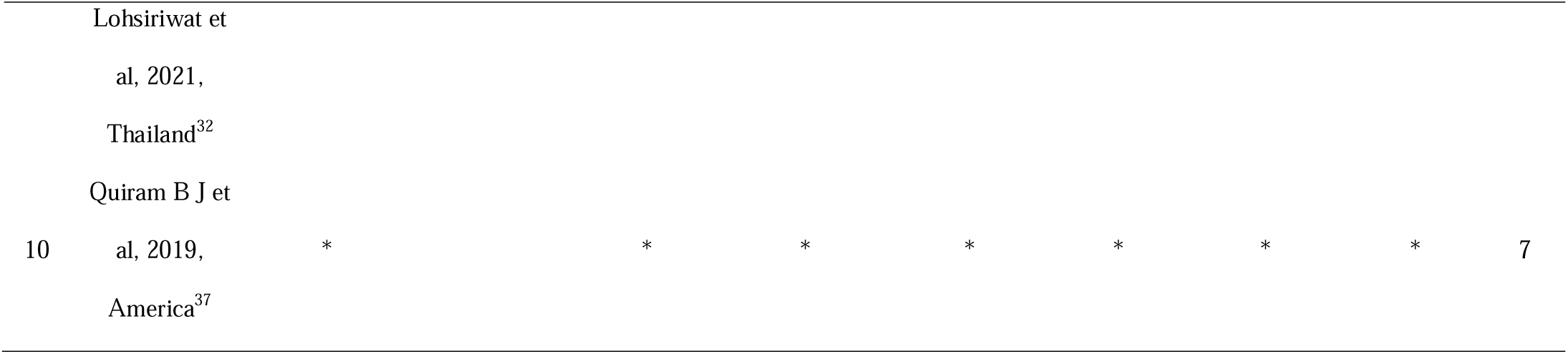
Newcastle-Ottawa Scale Quality Assessment Results for Included Studies Table 3 evaluates the quality of the 10 included studies using the Newcastle-Ottawa Scale (NOS), assessing three dimensions: “selection,” “comparability,” and “outcomes.” In the table, “*” indicates a 1-star rating for the corresponding item, while “**” denotes a 2-star rating. The total score ranges from 0 to 9 points, with higher scores indicating better study quality.

### Survival

#### KM-Based Meta-analysis

Data from 10 studies, including 5,867 postoperative patients with CRC^27–36^ were pooled to analyze long-term survival up to 10 years. During follow-up, 1,386 deaths occurred. The OS stratified by follow-up duration (1, 2, 3, 5 years) is shown in Figure 2: 1-year OS: 93.2% (95% CI: 92.3%-94.2%) in the ERAS group versus 91.5% (95% CI: 90.4%-92.6%) in controls; heterogeneity was low (I² = 0%, P = 0.68). A fixed-effect model showed significantly lower 1-year mortality in the ERAS group (odds ratio [OR] 0.67, 95% CI: 0.55-0.81, P < 0.001); 2-year OS: 86.7% (95% CI: 85.4%-88.0%) in the ERAS versus 84.3% (95% CI: 82.8%-85.8%) in controls; heterogeneity was low (I² = 0%, P = 0.44); fixed-effects OR 0.74 (95% CI: 0.64-0.85, P < 0.001); 3-year OS: 81.1% (95% CI: 79.6%-82.5%) versus 78.2% (95% CI: 76.5%-79.9%); I² = 44%, P = 0.07; fixed-effects OR 0.70 (95% CI: 0.61-0.79, P < 0.001); 5-year OS: 70.9% (95% CI: 69.0%-72.9%) versus 67.2% (95% CI: 65.1%-69.3%); I² = 62%, P = 0.01; random-effects OR 0.74 (95% CI: 0.59-0.93, P = 0.009 < 0.01). DFS (Figures 3) showed no significant mortality reduction, with 5-year OR 1.35 (95% CI: 1.10-1.67, P = 0.005 < 0.01).

**Figure 2:**
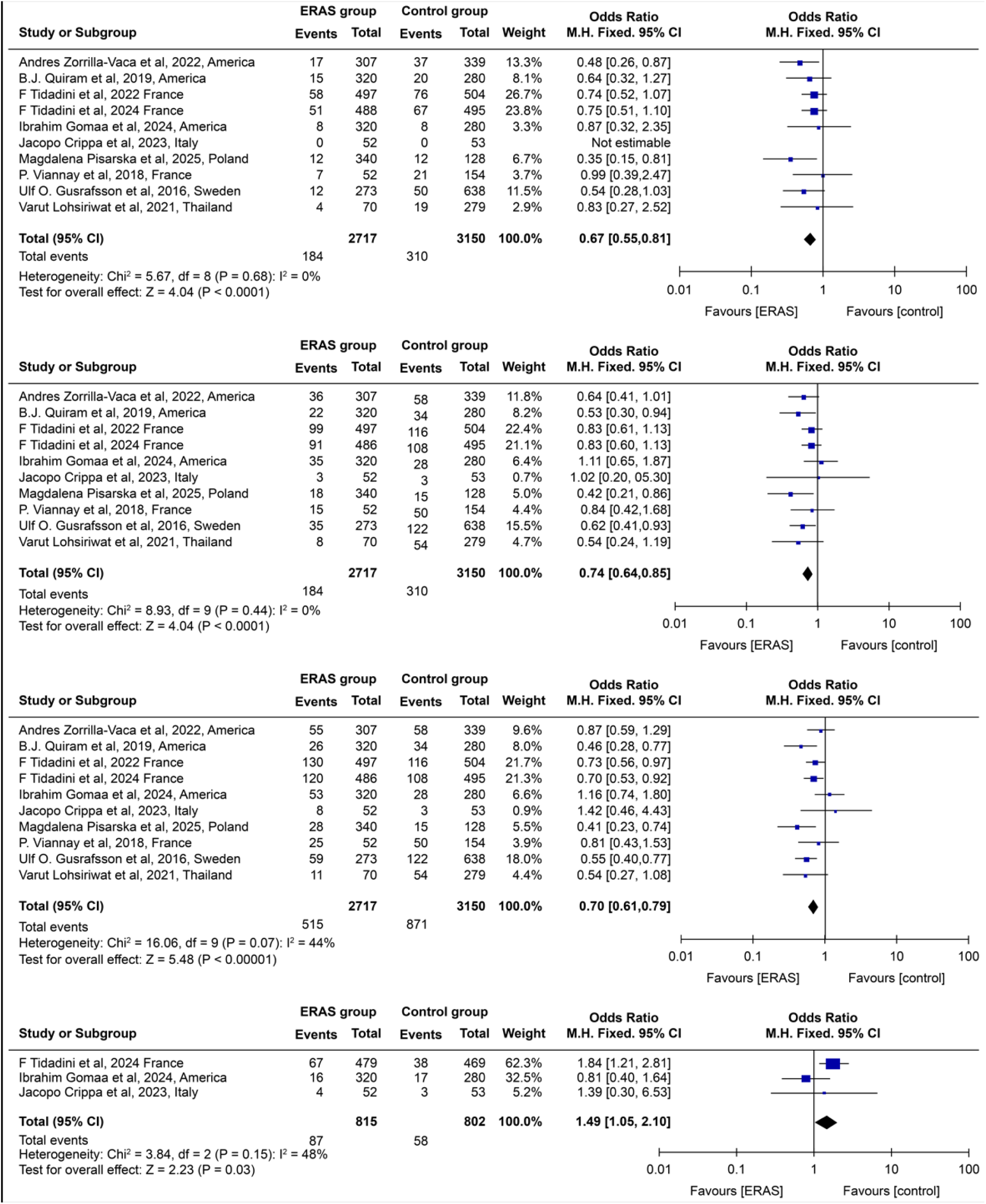
Meta-analysis forest plot of the impact of Enhanced Recovery After Surgery (ERAS) on overall survival (OS) at 1,2,3,5 years post-colorectal cancer surgery.

**Figure 3:**
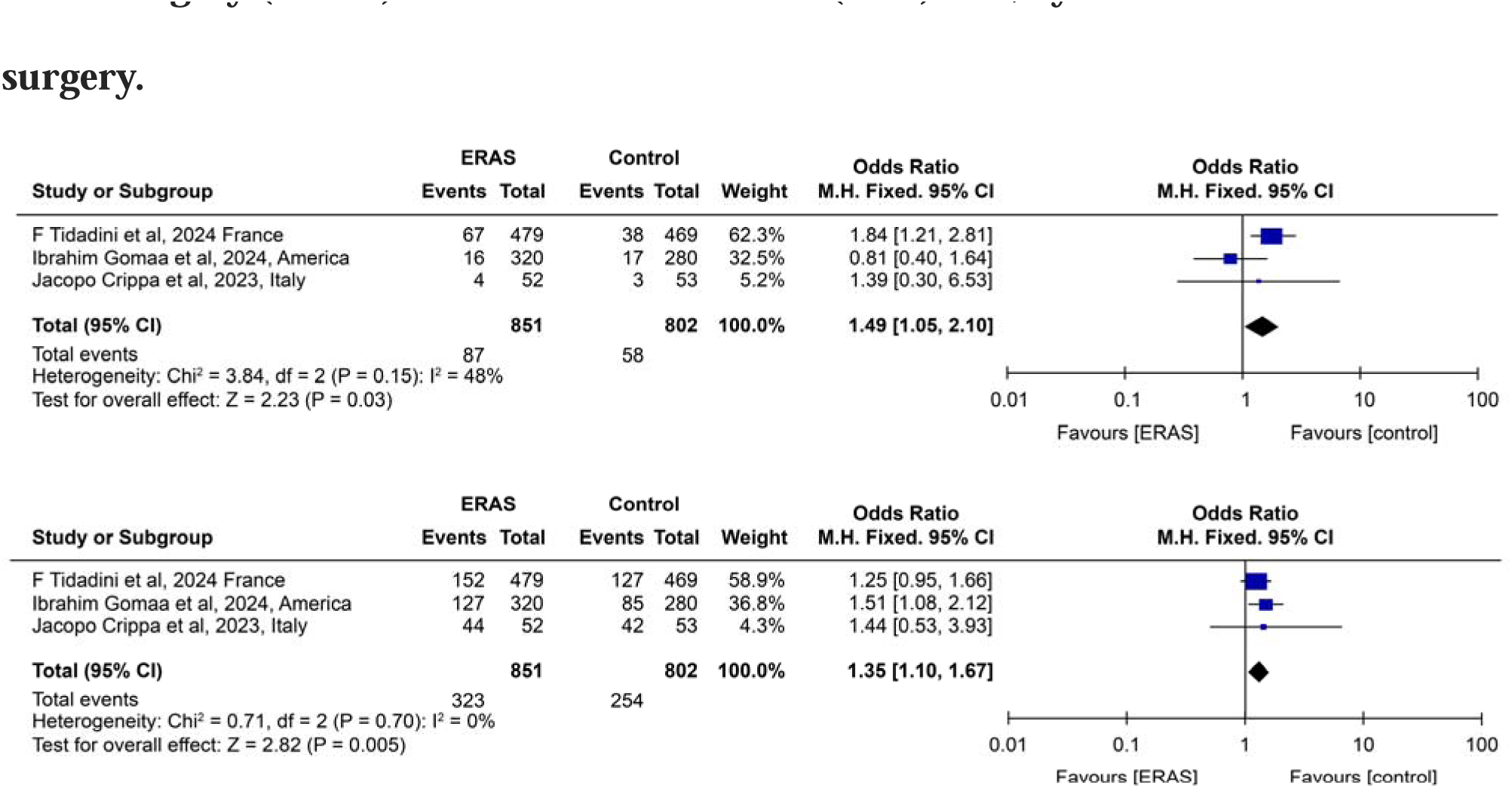
Meta-analysis forest plot of the effect of perioperative Enhanced Recovery After Surgery (ERAS) on disease-free survival (DFS) at 1,5 year after colorectal cancer surgery.

In summary, perioperative ERAS significantly improves 1-to 5-year OS in CRC, though it does not significantly affect long-term DFS.

#### Aggregated Data Meta-Analysis

Pooled data from eight studies (5,556 postoperative patients with CRC) evaluated the impact of ERAS on OS and DFS (Figure 4).^28,29,31,32,34–37^ All patients completed follow-up up to 10 years. OS analysis showed low heterogeneity (χ²=6.64, df=7, P=0.47; I²=0%), so a fixed-effect model was used. ERAS significantly reduced the long-term mortality risk (pooled risk ratio=0.72, 95% CI: 0.63-0.83; Z=4.76, P<0.00001).

**Figure 4:**
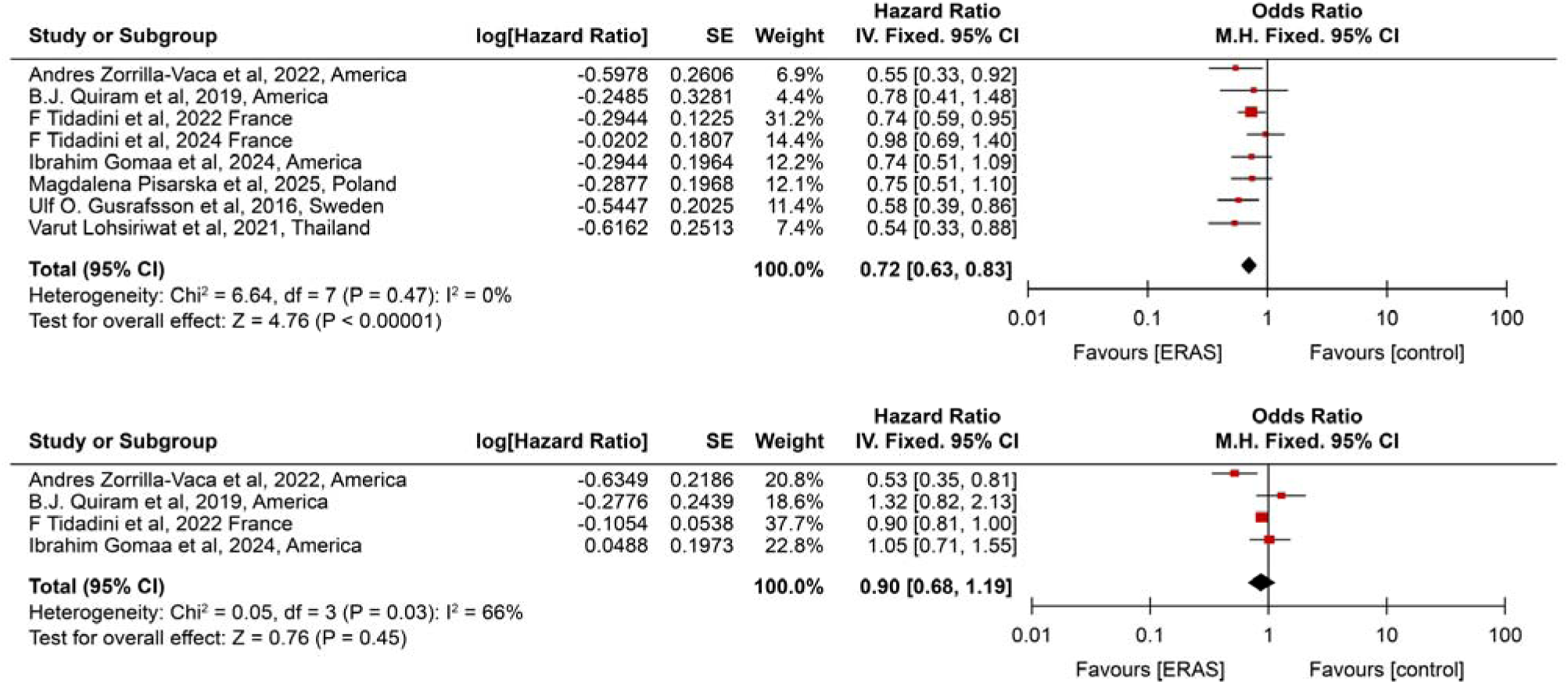
Meta-analysis forest plot of pooled data on the impact of Enhanced Recovery After Surgery (ERAS) on overall survival (OS) and disease-specific survival (DSF) following colorectal cancer surgery.

DFS analysis included four studies with significant heterogeneity (I²=66%), analyzed with a random-effects model. No significant difference in DFS was observed between ERAS and controls (pooled hazard ratio=0.90, 95% CI: 0.68-1.19; Z=0.76, P=0.45), indicating ERAS had no notable impact on DFS in patients with CRC.

#### Publication Bias Assessment

Funnel plots using 1-year (Supplement 2) and 3-year (Supplement 3) postoperative mortality ORs were constructed to assess publication bias. The scatter points were generally symmetric, with no apparent asymmetry or missing studies in the tails, indicating a low likelihood of publication bias in the meta-analysis.

### Subgroup Analysis

#### Effect Differences Across Tumor Stages

Data from five studies were pooled to evaluate the impact of perioperative ERAS on OS in patients with CRC by tumor stages (Stage I-II vs. Stage III-IV) (Figure 5).^29,31,32,35,37^ Heterogeneity was minimal (χ² = 2.02, df = 4, P = 0.73; I² = 0%), so a fixed-effect model was used. Stage I-II patients receiving ERAS had a significantly lower mortality risk than stage III-IV patients, with a pooled HR of 0.61 (95% CI: 0.50-0.73, Z=5.09, P<0.00001), indicating that patients with earlier-stage CRC derive greater survival benefit, with a more pronounced reduction in mortality risk, from perioperative ERAS.

**Figure 5:**
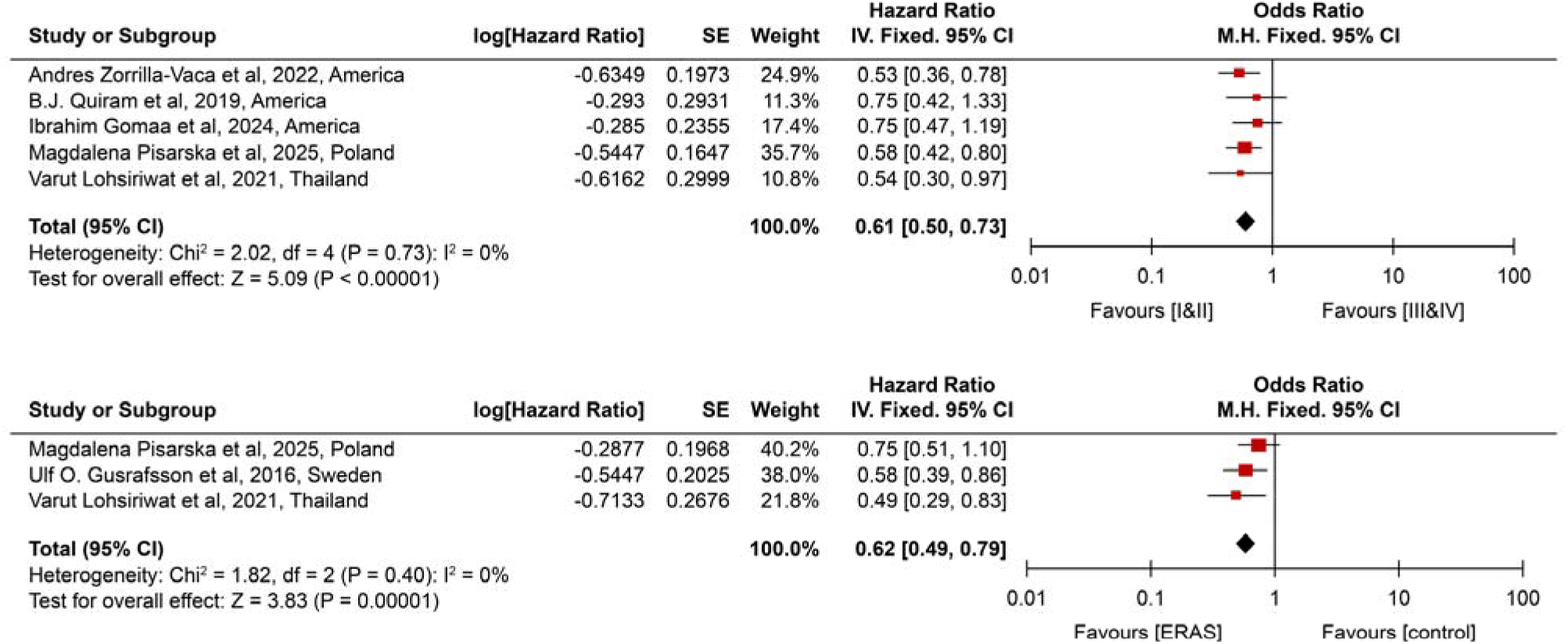
Meta-analysis forest plot of the impact of perioperative ERAS on overall survival in colorectal cancer patients with different tumor stages (Stage I–II vs. Stage III–IV) and varying ERAS compliance (high adherence vs. low adherence).

#### Effect of Different ERAS Adherence Levels

Data from three studies were pooled to evaluate OS in patients with CRC with high (≥70% or 80%) versus low adherence to perioperative ERAS protocols (Figure 5).^32,34,35^ Heterogeneity was low (χ² = 1.82, df = 2, P = 0.40; I² = 0%), so a fixed-effect model was applied. Patients with high adherence had a significantly lower mortality risk than those with low adherence, with a pooled HR of 0.62 (95% CI: 0.49-0.79; Z = 3.83, P = 0.0001), suggesting that greater adherence with ERAS is associated with reduced long-term mortality.

## Discussion

The ERAS concept was first introduced by Danish surgeon Dr. Kehlet in the 1990s. Its core principle lies in integrating evidence-based perioperative strategies that effectively mitigate surgical stress responses and promote rapid patient recovery.^38^ CRC surgery was among the first specialties to adopt ERAS. In 2005, the ERAS Society published the first guidelines specifically for CRC surgery, providing a structured framework for clinical practice.^39^ Extensive clinical evidence demonstrates that perioperative ERAS in patients with CRC significantly improves short-term outcomes, reduces postoperative complications, accelerates recovery, shortens hospital stays, and lowers healthcare costs.^4,5^ These demonstrated benefits have established a solid foundation for the continued and expanding application of ERAS in CRC care.

With the widespread adoption of ERAS and longer follow-up periods, its impact on long-term outcomes in patients with cancer has garnered considerable attention. F. Tidadini et al. examined 981 patients, comparing 5-year survival between an ERAS group and a conventional care group prior to ERAS implementation, and found no significant differences in OS or recurrence-free survival. In contrast, studies by Gomaa et al. reported that ERAS significantly improved long-term survival at 3 to 5 years postoperatively, highlighting inconsistencies in the literature.^28–31^ Through a systematic analysis of 10 studies encompassing 5,867 patients, this study provides robust evidence that perioperative ERAS significantly enhances 1-to 5-year OS in CRC, reducing long-term mortality risk by 28% (HR=0.72). This finding challenges the previous perception that ERAS primarily facilitates short-term recovery and confirms its capacity to significantly lower mortality risk at all evaluated time points (P<0.01), suggesting that ERAS delivers meaningful survival benefits within comprehensive cancer care.

The improvement in long-term survival associated with ERAS arises from synergistic effects across multiple domains, with core mechanisms focusing on three key dimensions: modulation of inflammation and immunity, prevention and management of complications, and continuity of care.^46–50^ Early postoperative feeding and mobilization further enhance recovery.^51^ By combining early nutrition, physical activity, and multimodal analgesia, ERAS protocols reduce postoperative hospital stays in patients with CRC by an average of 2.5 days and decrease overall complication risks by 48%.^52^

Additionally, ERAS promotes the timely and complete administration of adjuvant therapy. Delays or dose reductions in adjuvant therapy following cancer surgery are associated with lower 5-year survival, whereas ERAS accelerates recovery, enabling initiation of adjuvant therapy approximately 2-3 weeks earlier.^53,54^ A recent multicenter RCT integrating ERAS with electroacupuncture demonstrated a significant reduction in postoperative intestinal obstruction (POI) duration and a 49% lower risk of delayed POI.^55^

A comprehensive global ERAS framework is now established. The ERAS Society’s CRC guidelines comprise 22 core measures spanning preoperative, intraoperative, and postoperative management. Preoperative measures include patient education, nutritional screening and intervention, shortened fasting, and prophylactic analgesia. Intraoperative strategies encompass minimally invasive surgery, temperature management (operating room ≥21°C), goal-directed fluid therapy, and multimodal anesthesia. Postoperative measures include early oral feeding (within 24 h), early ambulation, minimized opioid use, and individualized drainage management.^56^ The completeness and quality of ERAS protocol implementation are critical to achieving long-term benefits.^57^

Despite clear standards, achieving high ERAS compliance in clinical practice remains challenging. Subgroup analyses show greater benefit in patients with stage I-II tumors and in those with high ERAS compliance. Several studies report overall ERAS compliance rates in CRC of only 50%-70%, with compliance to measures such as optimized bowel preparation and early mobilization often below 40%.^35,58,59^ In this meta-analysis, patients with high ERAS compliance (≥70% or 80%) had a 38% lower mortality risk (HR=0.62, 95% CI: 0.49-0.79) than those with low compliance, reinforcing compliance as a key determinant of ERAS effectiveness. Implementing standardized protocols and real-time monitoring systems may increase the proportion of high-compliance patients (≥70%) by over 30%.^57^

In this study, ERAS was associated with a significant improvement in DFS among patients with CRC, in contrast to the observed OS benefit. This discrepancy may be explained from three perspectives: First, the combined sample size of the three studies included in the DFS analysis was limited and significantly smaller than that of the OS analysis (5,867 cases). The small sample sizes reduced statistical power (62%), potentially limiting the ability to detect modest effects of ERAS on DFS.^60^

Second, most included studies were observational cohorts without rigorous adjustment or matching for key confounders, such as tumor histology and postoperative adjuvant therapy regimens, which may have diluted the independent effect of ERAS on DFS. Furthermore, variations in the definition of “disease-free survival” across studies resulted in inconsistent outcome assessment criteria, further weakening the observed association.^61^

Finally, the primary advantage of ERAS lies in enhancing surgical safety and completeness (e.g., reducing intraoperative injury and improving resection integrity), rather than directly preventing postoperative recurrence. This may partly explain the absence of significant differences in DFS. Although ERAS may exert indirect antitumor effects by attenuating surgical stress and preserving immune function, these benefits may manifest with a temporal lag.^62^

Overall, the lack of significant improvement in DFS is more likely attributable to methodological factors such as study design, heterogeneity in outcome definitions, and insufficient follow-up, rather than an inherent inability of ERAS to influence tumor recurrence. Future research should standardize DFS definitions, rigorously control confounding factors, and conduct large-scale, RCTs with long-term follow-up to clarify the true impact of ERAS on DFS in patients with CRC.

This study has certain limitations. First, most of the 10 included studies were cohort studies, with few high-quality RCTs, which may introduce selection bias. Second, there were differences in the specific implementation of ERAS measures and in compliance assessment criteria across studies. Although subgroup analyses reduced heterogeneity, residual bias may still remain. Finally, the included studies did not report the independent effects of individual ERAS components, limiting the ability to identify and prioritize the most beneficial measures.

Future research should prioritize multicenter, large-sample RCTs. Concurrently, investigations should be expanded by integrating advanced technologies such as single-cell sequencing and proteomics to further elucidate the microscopic signaling pathways involved in ERAS-mediated immune regulation and to identify molecular targets for optimizing ERAS protocols.

In conclusion, this study confirms that perioperative ERAS protocols in patients with CRC significantly improve 1-to 5-year survival rates and reduce mortality risk, with greater benefits observed in patients with early-stage tumors and high compliance. By refining implementation systems, enhancing compliance, and advancing mechanistic research, ERAS holds promise as a core component of comprehensive CRC care, delivering improved postoperative outcomes and long-term survival benefits for patients.

## Author Contributions

Ms Yang and Ms Liu had full access to all of the data in the study and take responsibility for the integrity of the data and the accuracy of the data analysis.

## Concept and design

Yang, Liu, Cui, Wu.

## Acquisition, analysis, or interpretation of data

All authors.

## Drafting of the manuscript

Yang, Liu, Cui, Wu

## Critical review of the manuscript for important intellectual content

All authors.

## Statistical analysis

Yang, Liu, Cui.

## Supervision

Liu, Wu.

## Conflict of Interest Disclosures

None reported.

## Data Sharing Statement

See Supplement 4.

## Additional Contributions

None

## Data Availability

All data produced in the present study are available upon reasonable request to the authors

## Notes

### Competing Interest Statement

The authors have declared no competing interest.

### Funding Statement

This study was funded by Shandong Provincial Clinical Key Specialty Discipline Development Funding

