## Supplement 1-3 for "Enhanced Recovery After Surgery (ERAS) Increases Long-Term Survival Rate after Surgery in Colorectal Cancer Patients: A Systematic Review and Meta-Analysis"

**1 Search Query:**

| **No.** | **Search Query** | **Number of Results** |
| --- | --- | --- |
| 1 | "Colorectal Neoplasms"[Mesh] | 259，193 |
| 2 | (((Colorectal Neoplasm[Title/Abstract]) OR (Neoplasm, Colorectal[Title/Abstract])) OR (Colorectal Tumors[Title/Abstract])) OR (Colorectal Tumor[Title/Abstract])) OR (Tumor, Colorectal[Title/Abstract]) OR (Tumors, Colorectal[Title/Abstract]) OR (Neoplasms, Colorectal[Title/Abstract]) OR (Colorectal Cancer[Title/Abstract])) OR (Cancer, Colorectal[Title/Abstract])) OR (Cancers, Colorectal[Title/Abstract])) OR (Colorectal Cancers[Title/Abstract])) OR (Colorectal Carcinoma[Title/Abstract])) OR (Carcinoma, Colorectal[Title/Abstract])) OR (Carcinomas, Colorectal[Title/Abstract])) OR (Colorectal Carcinomas[Title/Abstract]))) | 174，974 |
| 3 | (Enhanced Postsurgical Recovery) OR (Postsurgical Recoveries, Enhanced) OR (Postsurgical Recovery, Enhanced) OR (Recovery, Enhanced Postsurgical) OR ("Enhanced Recovery After Surgery"[Mesh]) | 10，440 |
| 4 | ("Survival Rate"[Mesh]) OR ( Overall Survival) OR ( long-term outcome) | 3,156,947 |
| 5 | ((Enhanced Postsurgical Recovery) OR (Postsurgical Recoveries, Enhanced) OR (Postsurgical Recovery, Enhanced) OR (Recovery, Enhanced Postsurgical) OR ("Enhanced Recovery After Surgery"[Mesh])) AND (("Survival Rate"[Mesh]) OR ( Overall Survival) OR ( long-term outcome)) | 1,994 |

**2: Publication bias funnel plot for 1-year postoperative mortality risk in colorectal cancer**

Plotted using the odds ratio (OR) for 1-year postoperative mortality risk as the indicator, the scatter points in the figure are broadly symmetrically distributed on both sides of the funnel, suggesting a low likelihood of publication bias in the meta-analysis for mortality risk at this time point.

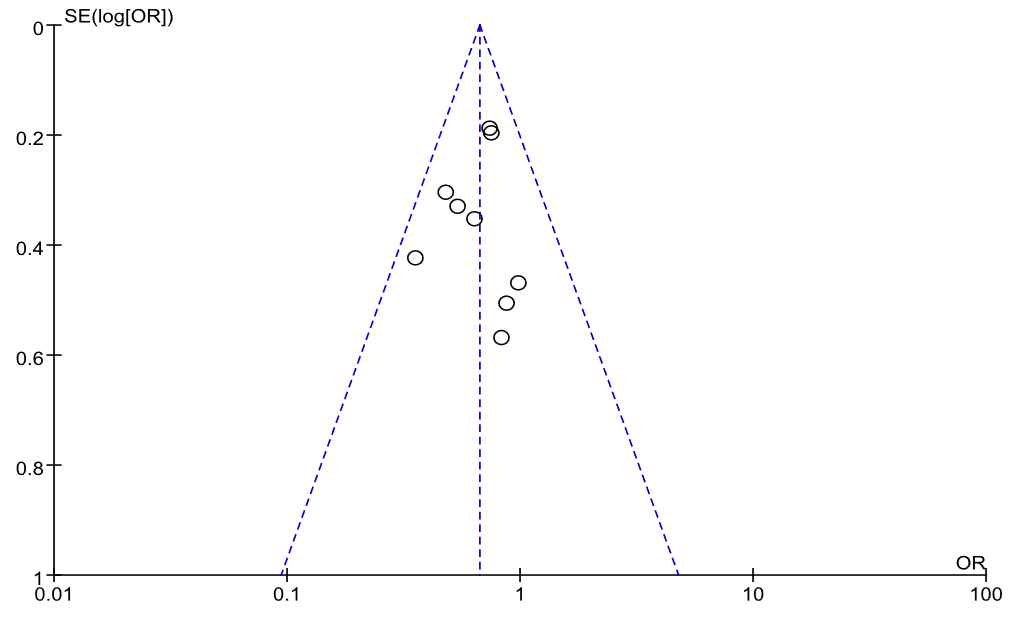

**3: Publication bias funnel plot for 3-year postoperative mortality risk in colorectal cancer**

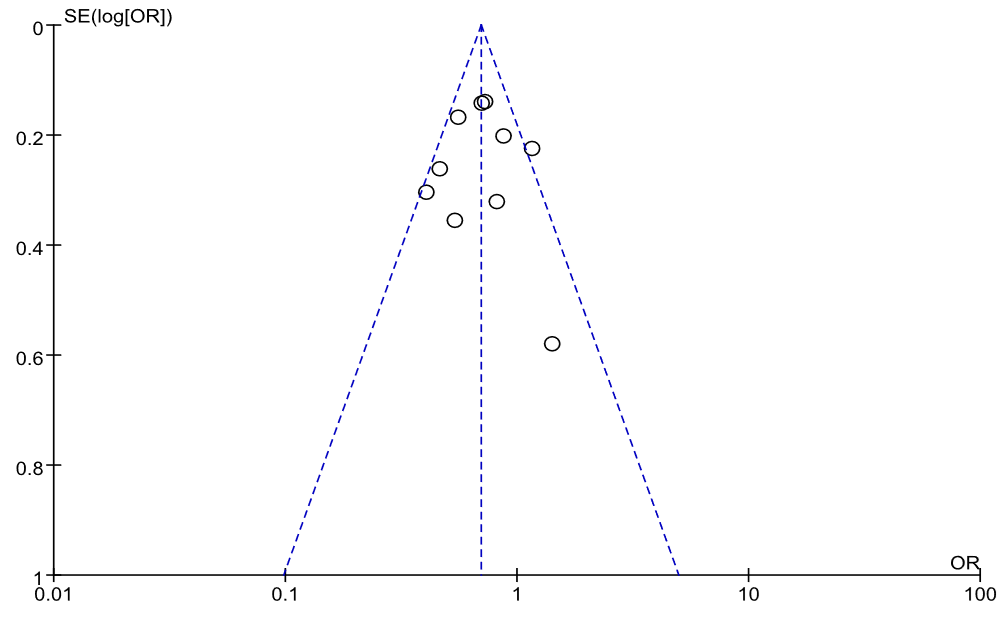
